# Maternal anaemia and polycythaemia during pregnancy and risk of inappropriate birthweight for-gestational-age babies: a retrospective cohort study in the northern belt of Ghana

**DOI:** 10.1101/2023.11.19.23298744

**Authors:** Silas Adjei-Gyamfi, Mary Sakina Zakaria, Abigail Asirifi, Issahaku Sulley, Mohammed Awal Ibrahim, Paul Armah Aryee

## Abstract

Small-for-gestational-age (SGA) and large-for-gestational-age (LGA) births are topical issues due to their devastating effects on the life course which are also accountable for neonatal mortalities and long-term morbidities.

**Objectives:** We tested the hypothesis that abnormal haemoglobin levels in each trimester of pregnancy will increase the risk of SGA and LGA deliveries in Northern Ghana.

**Design:** Retrospective cohort study was conducted from April to July 2020.

**Settings and Participants:** 422 postpartum mothers who had delivered within six weeks prior to the study were recruited through systematic random sampling from five primary and public health facilities in Northern Ghana.

**Primary outcome measure:** Using INTERGROWTH standards, SGA and LGA births were computed. Haemoglobin levels from antenatal records were analyzed to determine their effect on SGA and LGA births by employing multinomial logistic regression after adjusting for sociodemographic and obstetric factors at a significance level of α=0.05.

**Results:** Prevalence of anaemia in the first, second, and third trimesters of pregnancy was 63.5%, 71.3%, and 45.3% respectively and that of polycythaemia in the corresponding trimesters of pregnancy was 5.9%, 3.6%, and 1.7%. About 8.8% and 9.2% of the women delivered SGA and LGA babies respectively. After adjusting for confounders, anaemic mothers in the third trimester of pregnancy had increased risk of SGA births (aOR:5.56; 95%CI:0.64–48; p<0.001). Mothers with polycythaemia in the first, second, and third trimesters of pregnancy were 93% (aOR:0.07; 95%CI:0.01–0.46; p<0.040), 85% (aOR:0.15; 95%CI:0.08–1.65; p<0.001), and 88% (aOR:0.12; 95%CI:0.07–2.15; p=0.001) protected from SGA births respectively. Additionally, anaemia and polycythaemia across all trimesters of pregnancy were not statistically significant with LGA births.

**Conclusion:** The prevalence of anaemia in each trimester of pregnancy increased and that of polycythaemia decreased as the pregnancy progressed from first to third trimester. Delivery of LGA babies was more predominant compared to SGA babies. While anaemia in the third trimester of pregnancy increased the risk of SGA births, polycythaemia across the trimesters gave protection against SGA births. Healthcare providers and stakeholders should target pressing interventions for anaemia throughout pregnancy, especially during the third trimester.

**Article summary:** **Strengths and limitations of this study**

- Data employed in this analysis are from antenatal and/or delivery records of postpartum mothers attending postnatal care services in five major public health facilities in Savelugu municipality of Northern Ghana. Hence, the findings are generalizable to the populations in the entire municipality and its neighbouring districts.
- The measurement of anaemia and polycythaemia across all trimesters of pregnancy, small-for-gestational-age (SGA), and large-for-gestational-age (LGA) births were based on WHO and INTERGROWTH standards which gives an indication of objective assessment of the effect of these abnormal haemoglobin levels on SGA and LGA babies.
- This study provides vital evidence on the population-based effect of anaemia and polycythaemia across all trimesters of pregnancy on SGA and LGA babies among larger or several ethnic groups and broader age categories in the municipality.
- Data employed in the analysis were collected at a certain period of time (thus, April to July 2020), and also from antenatal records, so it could be difficult to draw causal inferences for the effect of anaemia and polycythaemia across all trimesters of pregnancy on SGA and LGA babies. Notwithstanding, the findings provide deep insights for health providers and stakeholders to identify prompt interventions for the prevention and treatment of abnormal haemoglobin levels.

## Introduction

In both developed and developing countries, low and high haemoglobin levels during pregnancy have been a topical issue (1–3) due to their devastating effects on birth outcomes and life course which subsequently leads to long-term health problems such as diabetes, cancer, hypertension, and stroke (4–7). Haemoglobin is an iron-containing oxygen transport protein in the erythrocytes of humans that carries oxygen from the lungs to different body parts (8,9). Haemoglobin level during pregnancy is categorized into low (haemoglobin levels less than 11g/dL), normal (haemoglobin levels from 11 to 13.1g/dL), and high or elevated (haemoglobin levels equal to or above 13.2g/dL) (2,9). The low and high haemoglobin levels are regarded as abnormal and low levels are termed as anaemia, while elevated levels are called polycythaemia (3,9). A measure of haemoglobin level (concentration) does not only serve as the best reliable and trustworthy indicator of anaemia and polycythaemia at the population level but also remains the standard test for pregnant women during antenatal visits which is used to assess anaemia or polycythaemia life (2,3). However, this measurement or test does not indicate the cause (9).

Haider and his colleagues emphasized that assessing low haemoglobin levels (anaemia) in pregnancy cannot be simply done through the measurement of blood haemoglobin concentration since anaemia rises more than fourfold from first to the third trimester (10). This proven fact of physiological fall is attributed to the expansion of plasma volume and reduction in the blood viscosity (11) which enhances the flow of blood in the low-pressure system of the intervillous space in the placenta for better foetal growth (12,13). Whilst elevated haemoglobin values in late pregnancy could mirror an inadequate expansion of blood plasma volume, low haemoglobin in early pregnancy might symbolize iron deficiency or other nutritional deficiencies including vitamins and folic acid (14). There is also a decrease in haemoglobin concentrations typically during the first trimester, reaching the lowermost concentration at the end of second trimester and then increasing again during the third trimester of pregnancy (15). Moreover, in women who are not given iron supplements, their haemoglobin levels fall sharply at 20 weeks gestation, then remain fairly constant up to 30 weeks gestation, and finally rises slightly afterward (16,17).

The global mean prevalence of haemoglobin in pregnancy is 11.4% with anaemia predominance of 38.2% (3). Gestational anaemia in Africa was estimated between 35.0% and 75.0% as compared to the developed countries where prevalence was found to be 15.0% (2,3,9). More than half of pregnancies found in Egypt (51.0%) (18), Northern Ghana (53.2%) (19), Kenya (57.0%) (20) and Southern Ethiopia (61.6%) (21) are anaemic which could be different from anaemia recorded across each stage of pregnancy. In a Chinese retrospective study, mothers in their first trimester of pregnancy have lower anaemia prevalence of 2.7% than those in their second and third trimesters with a prevalence of 14.7% and 16.6% correspondingly (22). In a cross-sectional study, anaemia in first, second, and third trimesters found among Boditii pregnant mothers in Northern Ethiopia was 7.8%, 45.4%, and 46.7% respectively (21). In Nigeria, the prevalence of anaemia in the first and third stage of gestation was approximated at 55.0% and 70.0% (23,24) while 40.8%, 37.0%, and 47.4% of Ghanaian (25), Bangladesh (26), and Tanzanian (27) mothers had low haemoglobin levels in the first, second and third trimesters of pregnancy respectively. Since polycythaemia has been misperceived as a sign of good nutrition, much attention has not been paid to it. Hence, most studies do not consider polycythaemia as a public health issue.

The ability of women to deliver babies with inappropriate birthweight for their gestational age is key in obstetric care due to its serious consequences on maternal and child health, although not much importance has been given to this category of adverse birth outcomes as compared to others such as low birthweight, macrosomia, preterm, and post-term newborns (1,5).

Inappropriate birthweight for gestational age babies can be classified into small-for-gestational-age or small-for-date (SGA) and large-for-gestational-age or large-for-date (LGA). The World Health Organization (WHO) defines LGA as birthweight above the ninetieth percentile of the gestational age in a given reference population while SGA refers to birthweight below the sex-specific tenth percentile of the gestational age (5,28). The burden of SGA births is high-pitched in the world. In 2012, an approximated 23 million infants were born with SGA in developing countries. Among these adverse birth outcomes, 1.5 million were delivered prematurely.

Southern Asia has the highest SGA prevalence of 34.0% followed by the sub-Saharan Africa (29). In Europe, 31.0% of neonates were born small for their gestational age (30). Comparatively, the respective incidence of SGA in Thailand (31), Brazil (32), Lithuania (33), and India (34) is 2.6%, 4.5%, 10.4%, and 11.9% whilst retrospective review in Africa reported SGA prevalence of 7.2%, 14.4%, and 16.6 % in Nigeria (5), Uganda (35) and Tanzania (36) correspondingly. Alternatively, 10.5% and 15.7% of Brazilian (32) and Thai (31) women delivered LGA babies respectively. Moreover, other studies estimated an overall occurrence of LGA to be 19.0% in China (37) and 11.0% in the United States of America (38).

Anaemia and polycythaemia (thus, abnormal haemoglobin levels) in pregnancy are very deleterious and may have tremendous effects on foetal and birth outcomes including SGA and LGA (39–41). Some earlier studies have produced contrasting and debatable reports on the effect of abnormal haemoglobin levels on SGA and LGA births. Anaemia in the first trimester of pregnancy increased the risk of SGA (14,42–44) and LGA (45,46) births in some meta-analysis, systematic reviews, and cross-sectional studies. Chinese women with low haemoglobin levels in the third stage of pregnancy were associated with lesser risks of SGA neonates as well as increased risks of LGA infants (47). Notwithstanding, some prospective studies in Northern Africa, and Europe revealed certainly no link between SGA and anaemia in the first (48–50), second (14), and third (14,51) trimesters of pregnancy correspondingly. Asynchronously, polycythaemia during the first (52), second (53), and third (43) trimesters was also found among mothers with SGA babies in Netherland, Sri Lanka, and Finland respectively. There was also no relationship between elevated haemoglobin levels greater than 13.2 g/dl in the final trimester of pregnancy and SGA (54) as well as LGA (43) infants.

Based on the reviews above, there exist some controversies between maternal abnormal haemoglobin levels in each trimester of pregnancy and these adverse birth outcomes. Hence, the assessment of haemoglobin levels in which trimester should be taken as standard is still unclear and this makes it very difficult to comprehend the most susceptible time for the foetus due to anaemia or polycythaemia in pregnancy. This highlights the need for geographic-specific studies on the association of abnormal haemoglobin levels with SGA and LGA births. As reported by Adjei-Gyamfi and his colleagues, the prevalence of anaemia in the first, second, and third trimesters of pregnancy in Savelugu municipal was 45.0%, 56.0%, and 44.0% respectively. The same study revealed a corresponding polycythaemia incidence of 5.4%, 1.1%, and 1.7% across the three trimesters of pregnancy (55). These prevalence rates of anaemia in each trimester of pregnancy are very alarming, while that of polycythaemia are significant. However, their effects on SGA and LGA babies in the municipality are not established. Moreover, there is data paucity on the relationship between abnormal haemoglobin levels in the different trimesters of pregnancy and SGA or LGA births among the Ghanaian population. In view of these precedents, we tested the hypothesis to find out whether anaemia and/or polycythaemia in each trimester of pregnancy will increase the risk of SGA and LGA births in the northern belt of Ghana.

## Materials and methods

### Study design

We conducted a retrospective cohort study on postpartum mothers who had delivered in the last six weeks in the northern belt of Ghana. This approach was employed to determine the association of abnormal haemoglobin levels with SGA and LGA babies.

### Study setting

The study was conducted in Savelugu municipality of Northern Region of Ghana. This peri-urban municipality has five major public health facilities including the municipal hospital and 21 community-based health planning services (CHPS) zones that provide maternal, neonatal, and child health care services to all Savelugu inhabitants and its environs. The total antenatal registrants and deliveries in 2020 were 2,160 and 3,974 respectively while the proportion of postnatal registrants was estimated at 99% (56).

### Study population

A population comprising all postpartum mothers who had delivered in the last six weeks prior to the study and attending postnatal care was recruited. Postpartum mothers with no live births and without antenatal and/or delivery records were excluded.

### Sample size and sampling procedure

Applying the Cochran (1977) formula (57), 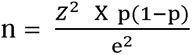 for sample size determination, the minimum sample size of 384 was initially approximated at 95% confidence interval corresponding to standard normal variate (z = 1.96), margin of error (e = 0.05), and a population proportion of 50% (p = 0.5) due to the paucity of available data on SGA and LGA births in Ghana. In order to cater for incomplete and damaged questionnaires, 10% of the sample was added to adjust the final sample size to 423.

The study included all five public health facilities in the municipality. In each health facility, the study sample was extracted using a systematic random sampling procedure. The names of the postpartum mother-child pairs in the postnatal care register were used as the sampling frame.

### Data collection

Based on existing literature and previous studies (47–50,55), a well-structured and pre-tested questionnaire was designed which was used to collect the data from April to July 2020 with the support of research assistants. Some of the data were collected from antenatal, delivery, and postnatal health records which included obstetric data (parity, gravidity, antenatal visits, mode of delivery, type of delivery, iron folic acid intake, sulphadoxine-pyrimethamine intake) and health status during pregnancy (malaria infection, tetanus-diphtheria immunization, family planning use, insecticide-treated bed nets use, haemoglobin levels at each trimester of pregnancy). Other maternal information were obtained through structured interview which were made up of socio-demographic characteristics (occupation, religion, age, education, ethnicity, socioeconomic status) and knowledge level on the association of abnormal haemoglobin with LGA and SGA babies.

### Measurement of study variables

The dependent variables were abnormal birthweight for gestational age (SGA and LGA) births while the principal independent variable was abnormal haemoglobin levels (anaemia and polycythaemia) in all trimesters of pregnancy.

LGA and SGA were assessed by using the INTERGROWTH-21st standard. LGA was defined and measured as birthweight above the ninetieth percentile of the gestational age in a given reference population (post-term newborns with large birthweight). Also, SGA is the birthweight below the tenth percentile of the gestational age (preterm newborns with low birthweight) (28,29,58). Haemoglobin level less than 11.0 g/dl at any of the trimesters of pregnancy was regarded as anaemia while haemoglobin level equal to or greater than 13.2g/dl was classified as polycythaemia (3,55).

The other independent variables of interest were collected and categorized based on previous studies, and biological plausibility (5,55). Mother’s knowledge level on the relationship of abnormal haemoglobin with SGA and LGA babies was summed up for each participant. Appropriate and inappropriate responses were scored one point and zero point respectively. Using the median cut-point, an absolute composite knowledge score was estimated using 48 items and categorized into adequate and inadequate knowledge levels, with a probable lowest score of zero and the highest score of 48 (55,59). Wealth index was assessed based on possession of household assets, housing quality, and availability of household utilities among others which were used as proxy indicators for the socioeconomic status (SES) of mothers. By using principal component analysis, the SES (wealth index) of the mothers was then trichotomized into high, middle, and low SES (60). Frequency of visits was categorized as less than eight visits and at least eight visits as per the WHO’s revised recommendations for positive pregnancy outcomes (61). Parity was grouped as one birth (delivery) and two or more births while gravidity was classified into one pregnancy and two or more pregnancies (55).

### Statistical analysis

The data set was cleaned and coded for statistical analysis using STATA 17.0 (Stata Corporation, Texas, USA). The dependent variable was coded as SGA = 1, appropriate-for-gestational-age (AGA) = 2, and LGA = 3. The “AGA” was used as the base outcome (reference point) during analyses.

Chi-square/Fisher’s exact tests were used to determine the association between dependent variables (SGA and LGA) and each independent variable. We further carried out univariate logistic regressions to show their strength of associations at a p-value of less than 0.05. Multinomial logistic regression was used to assess the association between SGA and LGA, and independent predictor variables while controlling for potential confounders. Some confounding variables adjusted for included maternal age, maternal knowledge level, antenatal visits, delivery mode, and sulphadoxine-pyrimethamine (SP) intake. Adjusted odds ratio (aOR) with a 95% confidence interval (CI) was run to identify statistically significant effect of abnormal haemoglobin levels (anaemia and polycythaemia) on SGA and LGA births.

### Ethical declarations

The Institutional Review Board of the Navrongo Health Research Centre offered ethical approval for this study (reference number: NHRCIRB373). The Savelugu Municipal Health Directorate granted permission to use the municipal health facilities as the study sites for data collection. Written informed consent/assent was obtained from all study participants and/or legal representatives before data collection.

## Results

### Background characteristics

Tables 1 and 2 describe the sociodemographic and obstetric characteristics of 422 women and baby pairs that were included in the study. However, one data was excluded from the analysis due to insufficient or incomplete information. The mean (sd) age of the studied women was 27.63 (6.02) years and that of the babies was 2.86 (1.97) weeks. Female (50.2%) and male (49.8%) babies were almost equally represented. Majority of the women were of Islamic religious group (88.2%), Dagomba/Mamprusi ethnic group (80.6%), and had marriage partners (92.2%). While a significant proportion of the women were self-employed (54.0%), about 43.1% of them had no formal education. 70.6% of the women had adequate knowledge on abnormal haemoglobin levels and/or their effects on SGA and LGA births. Of the studied participants, 73.7% of the women had two or more pregnancies and 71.6% of them had delivered two or more times. Surprisingly, less than one-third of the women (28.0%) made eight antenatal visits or more before delivery while pre-pregnancy underweight and overweight women were 3.1% and 30.8% respectively. Most of the women received iron-folic acid tablets (98.3%) and were immunized with tetanus-diphtheria (92.6%), while more than half of them took one to three doses of sulphadoxine-pyrimethamine (SP) (59.0%) during pregnancy. A greater number of the women delivered at health facilities (92.9%) and 89.8% had spontaneous vaginal delivery.

### Prevalence of anaemia and polycythaemia across trimesters of pregnancy and, small- and large-for-gestational-age births

The prevalence of anaemia in the first, second, and third trimesters of pregnancy was 63.5%, 71.3%, and 45.3% respectively. Additionally, the corresponding rate of polycythaemia in the first, second, and third trimesters of pregnancy was 5.9%, 3.6%, and 1.7% (Table 3). As shown in Figure 1, about 8.8% and 9.2% of the women delivered SGA and LGA babies respectively.

**Figure 1.**
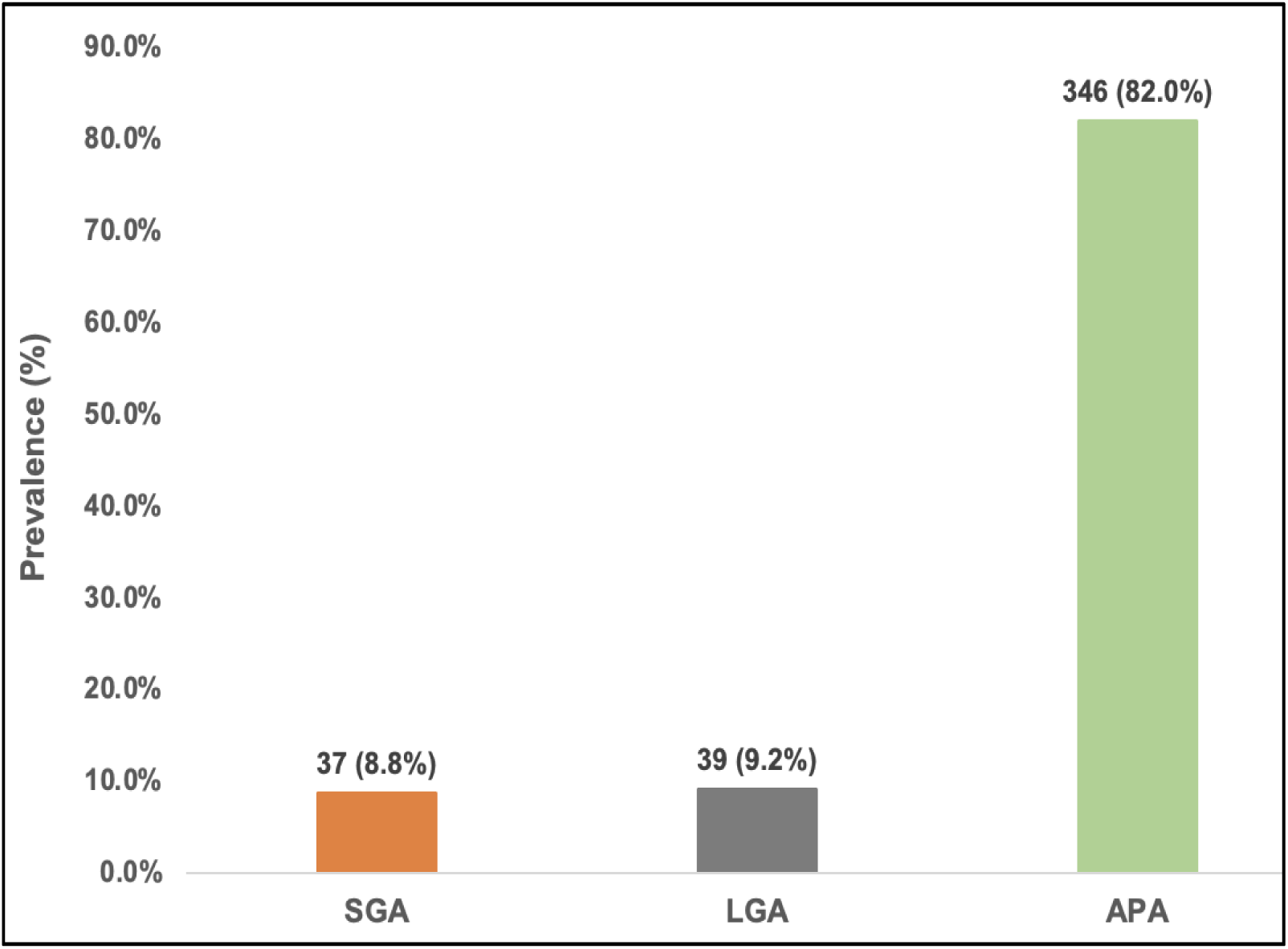
Prevalence of small- and large- for-gestational-age births.

### Bivariate association of small- and large-for-gestational-age births with background characteristics

At the bivariate level, maternal age (p=0.010), maternal knowledge (p=0.006), frequency of antenatal visits (p=0.006), SP intake (p=0.031), and delivery mode (p<0.001) were associated with abnormal size for gestational age (SGA and LGA) births (Tables 1 and 2). Additionally, anaemia in the second (p=0.014) and third (p<0.001) trimesters of pregnancy was significantly associated with SGA and LGA births. There was also significant association between polycythaemia in the first (p=0.002), second (p<0.001), and third (p=0.004) trimesters of pregnancy with SGA and LGA births (Table 3).

### Multivariate analysis for effect of anaemia and polycythaemia on small- and large-for-gestational-age births

The multivariate association between dependent and independent variables is exhibited in Table 4. After adjusting for background characteristics (maternal age, maternal knowledge, antenatal visits, SP intake, and delivery mode), anaemic mothers in the third trimester of pregnancy were 5.56 times more likely to deliver SGA babies (aOR:5.56; 95%CI:0.64–48; p<0.001). Mothers with polycythaemia in the first, second, and third trimesters of pregnancy reduced the risk of SGA births by 93% (aOR:0.07; 95%CI:0.01–0.46; p<0.040), 85% (aOR:0.15; 95%CI:0.08–1.65; p<0.001), and 88% (aOR:0.12; 95%CI:0.07–2.15; p=0.001) respectively. Moreover, teenage mothers (thus, mothers aged less than 20 years old) were more likely to give birth to SGA babies (aOR:4.47; 95%CI:1.51–13.3; p=0.007). Mothers with inadequate knowledge on abnormal haemoglobin levels and/or their effects on SGA and LGA births were about four times more likely to give birth to SGA babies (aOR:3.63; 95%CI:1.48–8.92; p=0.005).

Though there was no significant association between anaemia and polycythaemia in each trimester of pregnancy and LGA births after adjusting for maternal age, maternal knowledge level, antenatal visits, and SP intake, it was found that LGA babies were significantly associated with the likelihood of a mother delivering through CS mode (aOR:4.90; 95%CI:2.06–11.7; p<0.001).

## Discussion

Globally, abnormal haemoglobin levels and adverse birth outcomes like SGA and LGA births are recognized public health challenges principally found in developing countries which have unhealthy effects on adults’ life and are also responsible for the deleterious maternal and child health consequences including perinatal, child, and maternal mortalities (4,6). Attaining Sustainable Development Goal 3 by 2030 is a positive step to reduce these adverse birth outcomes (7). The study assessed the prevalence of anaemia and polycythaemia as well as SGA and LGA and explored the effects of these abnormal haemoglobin levels at different trimesters of pregnancy on SGA and LGA births in the Savelugu municipality of Northern region of Ghana.

The study reported mean (±sd) haemoglobin levels of 10.42 (±1.59) g/dl, 10.21 (±1.47) g/dl, and 10.68 (±1.56) g/dl in the first, second, and third trimesters of pregnancy respectively. The mean haemoglobin levels at each trimester in this study were lower than the findings from similar cohort respondents in Southern Ghana (49) and China (22). Thus, while the mean haemoglobin levels in each trimester of pregnancy in this study is classified as low (anaemia), that of the reported studies is categorized as normal. Anaemia in pregnancy is considered a serious and global public health challenge, especially in developing countries in Africa and Asia (9). The prevalence of anaemia in the first, second, and third trimesters of gestation in our study was estimated at 63.5%, 71.3%, and 45.2% respectively. Ghanaian data on anaemia and polycythaemia in each trimester of pregnancy is very limited (19,62). However, retrospective studies from Ethiopia (21) and China (22) respectively reported lower rates of anaemia prevalence in each trimester of pregnancy as compared to the present study. Anaemia prevalence in the three trimesters of pregnancy in this study is a critical public health concern since it is more than 40.0% of the population (3). The greater prevalence of anaemia in the study area could be a result of unhealthy practices such as pica (that is, an appetite deviation for non-food substances like ice, clay, soap, and chalk) commonly practiced among pregnant mothers in the Savelugu municipality (63). Therefore, the strong indication of anaemia among the pregnant populace in the municipality requires continuous and frequent nutrition education and counseling, especially on the intake of foods containing micronutrients like iron and folic acid (IFA) and/or vitamin B_12_ during pregnancy (64,65). There should also be a strict supply and intake of IFA supplements for pregnant mothers in the study area (66). Notwithstanding, the present study’s greater gestational anaemia in each trimester could also be due to different analyzer equipment for determination of haemoglobin concentration in the municipality. While the health centres use haemocue analyzers, the hospital, and private laboratories use haemotology analyzer machines which could bring about inconsistencies in detecting haemoglobin levels.

On the other hand, from the first to the third trimester of gestation, the present study disclosed the predominance of polycythaemia to be 5.9%, 3.6%, and 1.7% respectively. Thus, the polycythaemia status among the mothers reduces as the pregnancy progresses to term. Since polycythaemia among pregnant women has been misconceived as an affirmative indication of good nutrition worldwide, most studies do not report high haemoglobin levels in their findings. Nonetheless, appropriate education and counseling procedures must be rendered to mothers with polycythaemia due to its unwanted effects on pregnancy outcomes. Furthermore, advanced studies should consider polycythaemia as a public health concern and report on its prevalence s globally.

SGA prevalence in this study was 8.8% (95%CI: 6.2% – 11.9%). This finding is greater when compared with studies conducted in Thailand (31), and Brazil (32) with respective incidence rates of 2.6%, and 4.5%. Greater SGA prevalence in this study might be due to low antenatal attendance for routine check-ups and improved quality of care, as most mothers (72.0%) found in this study made less than eight antenatal visits. Lower prevalence of SGA was found in the present study as compared with previous reports from India (12.0%) (34) and Uganda (14.0%) (35). These previous studies (34,35) included more than 10 public and tertiary hospitals in their studies which might have resulted in a higher prevalence of SGA births.

Nonetheless, the present study estimated the prevalence of LGA to be 9.2% (95%CI: 6.7% – 12.4%), which was comparable with results from North America (USA) (38) and South America (Brazil) (32) with prevalence rates of 11.0% and 16.0% respectively. Pregnancy-related obesity and diabetes are very rampant among Americans (67) as compared to Ghanaians (19). Hence, pre-pregnancy obesity and gestational diabetes which are known predisposing factors for LGA births (32,67) might be the reason behind the greater LGA rates among these American mothers.

Our study findings showed that women with anaemia in the third trimester of pregnancy were more likely to give birth to SGA infants. In line with this study, many other study findings revealed significant relationship between haemoglobin levels lower than 11g/dL in the third trimester of pregnancy and SGA (68,69) while a retrospective inquiry from Pakistan was not consistent (51). Also, anaemia registered in the final stage of gestation among Chinese women was associated with lesser risks of SGA infants (47). Throughout pregnancy, there is a physiological decline in haemoglobin levels from early trimester to the third commonly projected at 5−14g/dl (16,70). This is attributable to the increase in blood plasma volume exceeding the rise in red cell mass (71) which results in the delivery of small-for-date babies. The final trimester of pregnancy is very significant as most environmental insults could affect the foetus serving as a critical window for the influence of anaemia on SGA deliveries (1,72). Anaemia during pregnancy could also be a sign of an underlying disease condition that can cause SGA births (72). Furthermore, of the 191 anaemic mothers found in the third trimester of pregnancy, the percentage of those who didn’t take more than 3 doses of SP (n = 143; 74.9%) is greater than those who ingested more than 3 doses (p < 0.001 in χ^2^/Fisher’s exact test). Greater proportion (n = 147; 77.0%) of these mothers with third-trimester anaemia attended less than eight ANC visits whilst very few made at least eight ANC visits (p = 0.040 in χ^2^/Fisher’s exact test). Thus, mothers who took few SP doses, and had lesser ANC visits during pregnancy increased the risk of developing anaemia (18,73,74) which could upsurge SGA births in the study area.

Maternal polycythaemia across all trimesters of pregnancy reduced the risk of SGA infants. This implies that mothers who had polycythaemia in each trimester of pregnancy served as significant protection against the delivery of SGA infants. Among the Northern Finland Birth Cohorts, maternal polycythaemia in the third trimester of gestation was statistically significant with an increased risk of SGA babies (43). Another study discovered an association between elevated haemoglobin levels and higher risk of small-weight babies among Welsh pregnant women in their second trimester (75). Additionally, in the Colombo district of Sri Lanka, polycythaemia during the second trimester of gestation was also found among mothers who delivered SGA infants (53). These previous studies are incongruent with the present study. Polycythaemia is associated with increased or adequate haemoglobin levels during pregnancy which supplies sufficient oxygen and nutrients to placenta and foetus leading to appropriate foetal growth and development which tends to be protective against SGA births. Further study is required to investigate the effects of elevated haemoglobin levels across each trimester of pregnancy on SGA births as well as other adverse birth outcomes in the municipal and entire Ghana.

After controlling for sociodemographic and obstetric variables, maternal age, and maternal knowledge were also associated with SGA births. Teenage mothers (thus, mothers aged less than 20 years) were more likely to give birth to SGA infants in the present study. An analogous pattern found among Tanzanian teenage mothers was associated with an increased risk of SGA babies (76). Our study is also similar to a cross-sectional study in Volta Ghana where mothers aged less than 20 were associated with small-for-date babies (77). Since teenage mothers are not physiologically, psychologically, and physically developed and matured enough to cope with the changing demands of pregnancy including foeto-maternal nutrition, and healthcare activities among others, it might lead to adverse birth outcomes like SGA births. Interestingly, the prevalence of teenage pregnancy in this study was 9.0% (95%CI: 6.5% – 12.2%). Though teenage pregnancy prevalence in our study is slightly lower than that of Ethiopia (12.0%) (78), it still calls for greater education and awareness on sexual and reproductive health with more emphasis on family planning methods because it places the municipality at risk of pregnancy-related complications among adolescents in the region.

In this study, less than half (n = 124; 29.4%) of mothers had inadequate knowledge on abnormal haemoglobin and/or its effects on SGA and LGA births which mirrored some cross-sectional studies in India and Kenya (79,80). The present study disclosed that having inadequate knowledge on abnormal haemoglobin levels and adverse birth outcomes increased the risk of SGA deliveries. The capacity of pregnant women to ascertain adequate knowledge on haemoglobin levels is very assistive and predictive in preventing and controlling adverse birth outcomes (79,81). In most Ghanaian society, women’s knowledge on the importance of haemoglobin levels may inform purchasing, supply, dietary and economic decisions. Therefore, mothers’ insufficient and/or acceptable awareness of the importance of haemoglobin levels may be associated with socio-cultural characteristics (82) which may probably lead to anaemia or polycythaemia during pregnancy (83) and could subsequently predict SGA births.

The results of the study showed that anaemia and polycythaemia in each trimester of pregnancy were not statistically associated with LGA births after adjusting for confounders. However, it was discovered that caesarian section (CS) mode of delivery was statistically associated with LGA births. CS delivery mode is a major consequence and competitive risk factor of large-weight births like LGA (84). Some African and Asian studies disclosed that mothers who delivered through CS mode were more likely to deliver LGA infants (33,85,86). Thus, these previous studies reflected the findings of our study. Notwithstanding LGA births are not regularly diagnosed and/or assessed in most Ghanaian health facilities. In developing countries, WHO recommends optimal CS delivery at a rate between 5.0% and 15.0% (87). The prevalence of CS delivery mode in this study was 10.2% (95%CI: 7.5% – 13.5%) which falls within the WHO recommendation. However, in developing countries, CS delivery poses a serious threat to maternal and child health due to the increased complications like excessive haemorrhage, vaginal tears, and asphyxiated neonates (88,89) leading to high incidence of maternal and perinatal deaths (90). Therefore, health service delivery should be well-equipped to quickly respond to CS delivery (91), especially in low-resource settings like Savelugu municipality. Obstetric tools such as the Friedman’s curve (also called WHO labour curve) should be applied and reinforced in all health facilities for reducing CS delivery rate. Additionally, the application of WHO non-clinical involvements such as childbirth preparation classes and frequent education on childbirth are commended to reduce CS deliveries (87) though they are influenced by the resource settings of a particular area.

### Limitations of the study

The prevalence of anaemia, polycythaemia, SGA, and LGA could be under or over-estimated by the time and/or season of data collection. This is because the data was collected from April to July 2020, which is the rainy and farming season for the Savelugu citizenry, so health visits are negatively affected. Hence, some respondents may not be available during data collection. Moreover, there might be possible sampling bias as a result of the non-selection of mothers who were absent from antenatal services in the first trimester of pregnancy or lost their antenatal records. Finally, information bias could occur since we collected some secondary data from the antenatal and/or delivery records. Some of the data might be incorrect due to inaccurate measurement, and documentation.

## Conclusion

The prevalence of anaemia across the trimesters of pregnancy was very high among the study participants which makes it a severe public health concern (3). In the descending order of prevalence, anaemia was highest in the second, followed by the first, and third trimesters of pregnancy. The prevalence of polycythaemia also decreased from the first trimester to term pregnancy. LGA babies were more predominant compared to SGA babies.

The study indicated that anaemia in the third trimester of pregnancy increased the risk of SGA births. Furthermore, mothers with polycythaemia across the trimesters of pregnancy had greater protection against SGA babies. Unpredictably, anaemia and polycythaemia in all the trimesters of pregnancy were not statistically significant with LGA births after adjusting for confounders.

Ghana Health Service (GHS) and other stakeholders should have a critical look at anaemia across all trimesters of pregnancy, especially during the third trimester. GHS should create greater awareness of third-trimester anaemia in pregnancy through collaboration with local governments to enhance public education on anaemia prevention in peri-urban communities and also take appropriate measures to treat it. GHS should sponsor in-service training activities to build the capacity of health professionals including midwives, community health nurses, and medical officers on the third-trimester anaemia in pregnancy and its effects on SGA births. This will help these cadres of health staff to copiously create awareness about the effects of third-trimester anaemia in pregnancy through health education via social media (WhatsApp, Tik tok, Instagram), leaflets, television, and radio within the municipality and beyond.

## Data Availability

The datasets will be made available by the corresponding author, without undue reservation. Kindly

## Declarations

### Patient and public involvement

Patients and/or the public were not involved in the design, or conduct, or reporting, or dissemination plans of this research.

### Patient consent for publication

Not applicable

### Consent for publication

None declared

### Availability of data and materials

The datasets will be made available by the corresponding author, without undue reservation. Kindly

### Competing interests

None declared

### Author statement

SAG, MSZ, AA, and PAA formulated the research questions, designed, and conceptualized the study. SAG, MSZ, AA, IS, and MAI carried out the data collection while SAG, IS, MAI, and PAA performed the data analysis, and interpretations. All authors contributed to reviewing and approving the final manuscript.

### Authors’ acronyms

Silas Adjei-Gyamfi (SAG); Mary Sakina Zakaria (MSK); Abigail Asirifi (AA); Issahaku Sulley (IS); Mohammed Awal Ibrahim (MAI); Paul Armah Aryee (PAA)

### Funding

This research received no specific grant from any funding agency in the public, commercial, or not-for-profit sectors.

## Acknowledgements

The authors wish to show their gratitude to the data collectors, study participants, and management of the study sites (health facilities) for their cooperation and unwavering support. We also thank Nutritional Science Department of University for Development Studies for offering supportive supervision during this study.

## List of abbreviations

ANC: Antenatal care
APA: Appropriate-for-gestational-age
APA: Appropriate-for-gestational-age
IFA: Iron and folic acid
LGA: Large-for-gestational-age
SGA: Small-for-gestational-age
SP: Sulphadoxine pyrimethamine
WHO: World Health Organization
USA: United States of America

## Notes

### Competing Interest Statement

The authors have declared no competing interest.

