## Supplemental Tables 1-4 for "Maternal anaemia and polycythaemia during pregnancy and risk of inappropriate birthweight for-gestational-age babies: a retrospective cohort study in the northern belt of Ghana"

**Table 1: Sociodemographic characteristics of respondents (n = 422)**

| **Characteristics** | **Frequency distribution** |  | **Small for gestational age** | **Appropriate for gestational age** | **Large for gestational age** | **p-value ^¶^** |
| --- | --- | --- | --- | --- | --- | --- |
|  | **Total (%)** |  | **SGA (%)** | **AGA (%)** | **LGA (%)** |  |
| **Maternal age group** |  |  |  |  |  |  |
| Less than 20 years | 38 (9.0) |  | 9 (24.3) | 27 (7.8) | 2 (5.1) | 0.010**^+^** |
| 20 – 35 years | 340 (80.6) |  | 26 (70.3) | 283 (81.8) | 31 (79.5) |  |
| More than 35 years | 44 (10.4) |  | 2 (5.4) | 36 (10.4) | 6 (15.4) |  |
| Mean (sd) = 27.63 (6.02) |  |  |  |  |  |  |
| **Marital status** |  |  |  |  |  |  |
| Single | 30 (7.1) |  | 6 (16.2) | 22 (6.4) | 2 (5.1) | 0.092 |
| Married | 389 (92.2) |  | 30 (81.1) | 322 (93.0) | 37 (94.9) |  |
| Divorced/widowed | 3 (0.7) |  | 1 (2.7) | 2 (0.6) | 0 (0.0) |  |
| **Maternal education status** |  |  |  |  |  |  |
| No formal | 182 (43.1) |  | 22 (59.5) | 197 (56.9) | 21 (53.8) | 0.884 |
| Formal | 240 (56.9) |  | 15 (40.5) | 149 (43.1) | 18 (46.2) |  |
| **Maternal ethnic group** |  |  |  |  |  |  |
| Gonja/Frafra | 56 (13.3) |  | 5 (13.5) | 42 (12.1) | 9 (23.1) | 0.356 |
| Dagomba/Mamprusi | 340 (80.6) |  | 28 (75.7) | 283 (81.8) | 29 (74.3) |  |
| Others ^&^ | 26 (6.1) |  | 4 (10.8) | 21 (6.1) | 1 (2.6) |  |
| **Religious affiliation** |  |  |  |  |  |  |
| Christianity | 49 (11.6) |  | 5 (13.5) | 38 (11.0) | 7 (18.0) | 0.710 |
| Islam | 373 (88.4) |  | 32 (86.5) | 308 (89.0) | 32 (82.0) |  |
| **Employment status** |  |  |  |  |  |  |
| Unemployed | 153 (36.3) |  | 11 (29.7) | 124 (35.8) | 18 (46.1) | 0.602 |
| Informal | 228 (54.0) |  | 22 (59.5) | 189 (54.6) | 17 (43.6) |  |
| Formal | 41 (9.7) |  | 4 (10.8) | 33 (9.6) | 4 (10.3) |  |
| **Maternal socioeconomic status** | |  |  |  |  |  |
| Low | 170 (40.3) |  | 15 (40.5) | 138 (39.9) | 17 (43.6) | 0.996 |
| Middle | 84 (19.9) |  | 7 (19.0) | 70 (20.2) | 7 (19.9) |  |
| High | 168 (39.8) |  | 15 (40.5) | 138 (39.9) | 15 (38.5) |  |
| **Sex of neonate** |  |  |  |  |  |  |
| Male | 210 (49.8) |  | 20 (54.0) | 172 (49.7) | 18 (46.2) | 0.788 |
| Female | 212 (50.2) |  | 17 (46.0) | 174 (50.3) | 21 (53.8) |  |
| **Maternal knowledge** |  |  |  |  |  |  |
| Inadequate | 124 (29.4) |  | 19 (51.4) | 97 (28.0) | 8 (20.5) | 0.006**^+^** |
| Adequate | 298 (70.6) |  | 18 (48.6) | 249 (72.0) | 31 (79.5) |  |

**^+^** p-value < 0.05	**^¶^** Chi-square/Fischer’ exact test	^&^ Asante, Ewe, Kusasi

**Table 2: Obstetric characteristics of respondents (n = 422)**

| **Characteristics** | **Frequency distribution** |  | **Small for gestational age** | **Appropriate for gestational age** | **Large for gestational age** | **p-value ^¶^** |
| --- | --- | --- | --- | --- | --- | --- |
|  | **Total (%)** |  | **SGA (%)** | **AGA (%)** | **LGA (%)** |  |
| **Number of pregnancies** |  |  |  |  |  |  |
| 0 – 1 | 111 (26.3) |  | 13 (35.1) | 92 (26.6) | 6 (15.4) | 0.142 |
| 2 or more | 311 (73.7) |  | 24 (64.9) | 254 (73.4) | 33 (84.6) |  |
| **Number of deliveries** |  |  |  |  |  |  |
| 0 – 1 | 120 (26.3) |  | 13 (35.1) | 100 (28.9) | 7 (18.0) | 0.228 |
| 2 or more | 302 (71.6) |  | 24 (64.9) | 246 (71.1) | 32 (82.0) |  |
| **Frequency of antenatal visits** | |  |  |  |  |  |
| Less than 8 | 304 (72.0) |  | 33 (89.2) | 249 (72.0) | 22 (56.4) | 0.006**^+^** |
| 8 or more | 118 (28.0) |  | 4 (10.8) | 97 (28.0) | 17 (43.6) |  |
| **Pre-pregnancy body mass index** | |  |  |  |  |  |
| Overweight | 130 (30.8) |  | 8 (21.6) | 106 (30.6) | 16 (41.0) | 0.438 |
| Normal | 279 (66.1) |  | 28 (75.7) | 229 (66.2) | 22 (56.4) |  |
| Underweight | 13 (3.1) |  | 1 (2.7) | 11 (3.2) | 1 (2.6) |  |
| **Sulphadoxine-pyrimethamine intake** | | | |  |  |  |
| None | 25 (5.9) |  | 2 (5.4) | 21 (6.1) | 2 (5.1) | 0.031**^+^** |
| 1 – 3 doses | 249 (59.0) |  | 28 (75.7) | 205 (59.2) | 16 (41.0) |  |
| > 3 doses | 148 (35.1) |  | 7 (18.9) | 120 (34.7) | 21 (53.9) |  |
| **Tetanus-diphtheria immunization** | |  |  |  |  |  |
| Not immunized | 31 (7.4) |  | 2 (5.4) | 25 (7.2) | 4 (10.3) | 0.706 |
| Immunized | 391 (92.6) |  | 35 (94.6) | 321 (92.8) | 35 (89.7) |  |
| **Insecticide-treated bed nets (ITNs) use** | | |  |  |  |  |
| No ITNs use | 152 (36.0) |  | 20 (54.0) | 119 (34.5) | 13 (33.3) | 0.057 |
| ITNs use | 270 (64.0) |  | 17 (46.0) | 227 (65.6) | 26 (66.7) |  |
| **Family planning (FP) use** | |  |  |  |  |  |
| No FP use | 347 (82.2) |  | 31 (83.8) | 286 (82.7) | 30 (76.9) | 0.652 |
| FP use | 75 (17.8) |  | 6 (16.2) | 60 (17.3) | 9 (23.1) |  |
| **Iron folic acid intake** |  |  |  |  |  |  |
| No | 7 (1.7) |  | 0 (0.0) | 7 (2.0) | 0 (0.0) | 1.000 |
| Yes | 415 (98.3) |  | 37 (100.0) | 339 (98.0) | 39 (100.0) |  |
| **Gestational malaria infection** | |  |  |  |  |  |
| Episode | 59 (14.0) |  | 32 (86.5) | 46 (13.3) | 8 (20.5) | 0.466 |
| No episode | 363 (86.0) |  | 5 (13.5) | 300 (86.7) | 31 (79.5) |  |
| **Type (place) of delivery** |  |  |  |  |  |  |
| Skilled (facility) | 392 (92.9) |  | 33 (89.2) | 325 (93.9) | 34 (87.2) | 0.196 |
| Unskilled (home) | 30 (7.1) |  | 4 (10.8) | 21 (6.1) | 5 (12.8) |  |
| **Mode of delivery** |  |  |  |  |  |  |
| Vaginal | 379 (89.8) |  | 31 (83.8) | 321 (92.8) | 27 (69.2) | < 0.001**^+^** |
| Caesarian section | 43 (10.2) |  | 6 (16.2) | 25 (7.2) | 12 (30.8) |  |

**^+^** p-value < 0.05	**^¶^** Chi-square/Fischer’ exact test

**Table 3: Association of anaemia and polycythaemia with small- and large-for-gestational-age births (n = 422)**

| **Haemoglobin (Hb) levels across trimesters of pregnancy** | **Prevalence** |  | **Small for gestational age** | **Appropriate for gestational age** | **Large for gestational age** | **p-value ^¶^** |
| --- | --- | --- | --- | --- | --- | --- |
|  | **Total (%)** |  | **n (%)** | **n (%)** | **n (%)** |  |
| **First-trimester Hb levels** |  |  |  |  |  |  |
| Anaemia (< 11 g/dl) | 268 (63.5) |  | 25 (67.6) | 221 (63.9) | 22 (56.4) | 0.568 |
| Normal (11 – 13.1 g/dl) | 129 (30.6) |  | 6 (16.2) | 111 (32.1) | 12 (30.8) | Reference |
| Polycythaemia (≥ 13.2) | 25 (5.9) |  | 6 (16.2) | 14 (4.0) | 5 (12.8) | 0.002**^+^** |
| **Second-trimester Hb levels** | |  |  |  |  |  |
| Anaemia | 301 (71.3) |  | 28 (75.7) | 253 (73.1) | 20 (51.3) | 0.014**^+^** |
| Normal | 106 (25.1) |  | 1 (2.7) | 88 (25.4) | 17 (43.6) | Reference |
| Polycythaemia | 15 (3.6) |  | 8 (21.6) | 5 (1.5) | 2 (5.1) | < 0.001**^+^** |
| **Third-trimester Hb levels** | |  |  |  |  |  |
| Anaemia | 191 (45.3) |  | 33 (89.2) | 147 (42.5) | 11 (28.2) | < 0.001**^+^** |
| Normal | 224 (53.0) |  | 1 (2.7) | 196 (56.6) | 27 (69.2) | Reference |
| Polycythaemia | 7 (1.7) |  | 3 (8.1) | 3 (0.9) | 1 (2.6) | 0.004**^+^** |

**^+^** p-value < 0.05	**^¶^** Chi-square/Fischer’ exact test

**Table 4: Multivariate analysis for effect of anaemia and polycythaemia on small- and large-for-gestational-age births (n = 422)**

| **Characteristics** |  | **Multinomial logistic regression (*Appropriate for gestational age = base outcome*)** | | | | | | | | |
| --- | --- | --- | --- | --- | --- | --- | --- | --- | --- | --- |
|  | | **SGA: Small-for-gestational-age** | | | |  | **LGA: Large-for-gestational-age** | | | |
|  |  | **cOR (95%CI)** | **p-value** | **aOR (95%CI)** | **p-value** |  | **cOR (95%CI)** | **p-value** | **aOR (95%CI)** | **p-value** |
| **First-trimester Hb levels** |  |  |  |  |  |  |  |  |  |  |
| Anaemia |  | 2.09 (0.83 – 5.25) | 0.116 | 1.91 (0.52 – 7.06) | 0.329 |  | 0.92 (0.44 – 1.92) | 0.827 | 1.27 (0.57 – 2.82) | 0.554 |
| Normal |  | Reference | | | |  | Reference | | | |
| Polycythaemia |  | 0.08 (0.02 – 0.28) | 0.001**^+^** | **0.07 (0.01 – 0.46)** | **0.040^+^** |  | 3.30 (1.01 – 10.8) | 0.048**^+^** | 2.90 (0.73 – 11.57) | 0.132 |
| **Second-trimester Hb levels** | |  |  |  |  |  |  |  |  |  |
| Anaemia |  | 9.74 (1.30 – 72.6) | 0.026**^+^** | 5.20 (0.63 – 42.8) | 0.125 |  | 0.41 (0.21 – 0.82) | 0.011**^+^** | 0.54 (0.25 – 1.15) | 0.109 |
| Normal |  | Reference | | | |  | Reference | | | |
| Polycythaemia |  | 0.14 (0.01 – 1.36) | < 0.001**^+^** | **0.15 (0.08 – 1.65)** | **< 0.001^+^** |  | 2.07 (0.37 – 11.6) | 0.407 | 1.33 (0.18 – 9.79) | 0.777 |
| **Third-trimester Hb levels** | |  |  |  |  |  |  |  |  |  |
| Anaemia |  | 4.40 (5.95 – 32.5) | < 0.001**^+^** | **5.56 (0.64 – 48.1)** | **< 0.001^+^** |  | 0.54 (0.26 – 1.13) | 0.103 | 0.75 (0.34 – 1.67) | 0.480 |
| Normal |  | Reference | | | |  | Reference | | | |
| Polycythaemia |  | 0.12 (0.02 – 2.47) | < 0.001**^+^** | **0.12 (0.07 – 2.15)** | **0.001^+^** |  | 2.42 (0.24 – 24.1) | 0.451 | 1.84 (0.15 – 22.8) | 0.634 |
| **Maternal age group** |  |  |  |  |  |  |  |  |  |  |
| Less than 20 years |  | 3.63 (1.54 – 8.53) | 0.003**^+^** | **4.47 (1.51 – 13.3)** | **0.007^+^** |  | 0.68 (0.15 – 2.98) | 0.605 | 0.87 (0.18 – 4.21) | 0.866 |
| 20 – 35 years |  | Reference | | | |  | Reference | | | |
| More than 35 years |  | 0.60 (0.14 – 2.65) | 0.505 | 0.29 (0.03 – 2.52) | 0.264 |  | 1.52 (0.59 – 3.90) | 0.382 | 1.74 (0.61 – 4.97) | 0.300 |
| **Maternal knowledge level** | | |  |  |  |  |  |  |  |  |
| Inadequate |  | 2.71 (1.36 – 5.38) | 0.004**^+^** | **3.63 (1.48 – 8.92)** | **0.005^+^** |  | 0.66 (0.29 – 1.49) | 0.320 | 0.78 (0.33 – 1.88) | 0.582 |
| Adequate |  | Reference | | | |  |  |  | Reference | |
| **Frequency of antenatal visits** | | |  |  |  |  |  |  |  |  |
| Less than 8 |  | 3.21 (1.11 – 9.31) | 0.031**^+^** | 1.94 (0.50 – 7.49) | 0.336 |  | 0.50 (0.26 – 0.99) | 0.047**^+^** | 0.69 (0.32 – 1.48) | 0.345 |
| 8 or more |  | Reference | | | |  | Reference | | | |
| **Sulphadoxine-pyrimethamine intake** | | |  |  |  |  |  |  |  |  |
| None |  | 0.70 (0.16 – 3.13) | 0.638 | 1.20 (0.23 – 6.37) | 0.830 |  | 1.22 (0.26 – 5.68) | 0.800 | 1.74 (0.34 – 8.82) | 0.502 |
| 1 – 3 doses |  | Reference | | | |  | Reference | | | |
| > 3 doses |  | 0.43 (0.18 – 1.01) | 0.052 | 0.73 (0.24 – 2.16) | 0.564 |  | 2.24 (1.13 – 4.46) | 0.021**^+^** | 1.94 (0.91 – 4.15) | 0.088 |
| **Mode of delivery** |  |  |  |  |  |  |  |  |  |  |
| Caesarian section |  | 2.49 (0.95 – 6.52) | 0.064 | 4.24 (1.03 – 17.4) | 0.065 |  | 5.71 (2.58 – 12.6) | < 0.001**^+^** | **4.90 (2.06 – 11.68)** | **< 0.001^+^** |
| Vaginal |  | Reference | | | |  | Reference | | | |
| ***Regression model*** |  |  |  |  |  |  |  |  |  |  |
| *R^2^* |  | *0.469* |  |  |  |  |  |  |  |  |
| *p-value* |  | *< 0.001****^+^*** |  |  |  |  |  |  |  |  |

**^+^** p-value < 0.05	*aOR: Adjusted odds ratio	cOR: Crude odds ratio*

| **Characteristics** | **Frequency distribution** |  | **LBW: Low birthweight** | | |  | **SGA: Small for gestational age** | | |
| --- | --- | --- | --- | --- | --- | --- | --- | --- | --- |
|  | **Total (%)**  **n = 422** |  | **LBW (%)**  **n = 55** | **No LBW (%)**  **n = 367** | **p-value ^a^** |  | **SGA (%)**  **n = 37** | **No SGA (%)**  **n = 385** | **p-value ^a^** |
| **Age group of mother** |  |  |  |  |  |  |  |  |  |
| < 20 years | 38 (9.0) |  | 11 (20.0) | 27 (7.4) | 0.002^*^ |  | 9 (24.3) | 29 (7.5) | 0.001^*^ |
| 20 – 35years | 340 (80.6) |  | 42 (76.4) | 298 (81.2) | Ref |  | 26 (70.3) | 314 (81.6) | Ref |
| > 35 years | 44 (10.4) |  | 2 (3.6) | 42 (11.4) | 0.077 |  | 2 (5.4) | 42 (10.9) | 0.295 |
| Mean (SD) = 27.63 (6.02) |  |  |  |  |  |  |  |  |  |
| **Marital status of mother** |  |  |  |  |  |  |  |  |  |
| Single | 3 (7.1) |  | 6 (10.9) | 24 (6.5) | 0.240 |  | 6 (16.2) | 24 (6.2) | 0.024^*^ |
| Married | 389 (92.2) |  | 47 (85.5) | 342 (93.2) | Ref |  | 30 (81.1) | 359 (93.3) | Ref |
| Divorced/widowed | 3 (0.7) |  | 2 (3.6) | 1 (0.3) | 0.006^*^ |  | 1 (2.7) | 2 (0.5) | 0.241 |
| **Education level of mother** |  |  |  |  |  |  |  |  |  |
| No formal | 182 (43.1) |  | 20 (36.4) | 162 (44.1) | 0.277 |  | 15 (40.5) | 167 (43.4) | 0.739 |
| Formal | 240 (56.9) |  | 35 (63.6) | 205 (55.9) | Ref |  | 22 (59.5) | 218 (56.6) | Ref |
| **Ethnicity of mother** |  |  |  |  |  |  |  |  |  |
| Gonja/Frafra | 56 (13.3) |  | 7 (12.7) | 49 (13.4) | 0.899 |  | 5 (13.5) | 51 (13.3) | 0.964 |
| Dagomba/Mamprusi | 340 (80.6) |  | 43 (78.2) | 297 (80.9) | Ref |  | 28 (75.7) | 312 (81.0) | Ref |
| Others | 26 (6.1) |  | 5 (9.1) | 21 (5.7) | 0.333 |  | 4 (10.8) | 22 (5.7) | 0.218 |
| **Religion of mother** |  |  |  |  |  |  |  |  |  |
| Christianity | 49 (11.6) |  | 8 (14.6) | 41 (11.2) | 0.715 |  | 5 (13.5) | 44 (11.4) | 0.889 |
| Islam | 373 (88.4) |  | 47 (85.4) | 326 (88.8) | Ref |  | 32 (86.5) | 341 (88.6) | Ref |
| **Occupation of mother** |  |  |  |  |  |  |  |  |  |
| Unemployed | 153 (36.3) |  | 17 (30.9) | 136 (37.1) | 0.376 |  | 11 (29.7) | 142 (36.9) | 0.387 |
| Informal | 228 (54.0) |  | 33 (60.0) | 195 (53.1) | Ref |  | 22 (59.5) | 206 (53.5) | Ref |
| Formal | 41 (9.7) |  | 5 (9.1) | 36 (9.8) | 0.867 |  | 4 (10.8) | 37 (9.6) | 0.814 |
| **Socioeconomic status of mother** |  |  |  |  |  |  |  |  |  |
| Low | 170 (40.3) |  | 23 (41.8) | 147 (40.1) | 0.804 |  | 15 (40.5) | 155 (40.3) | 0.973 |
| Middle | 84 (19.9) |  | 9 (16.4) | 75 (20.4) | 0.481 |  | 7 (19.0) | 77 (20.0) | 0.875 |
| High | 168 (39.8) |  | 23 (41.8) | 145 (39.5) | Ref |  | 15 (40.5) | 153 (39.7) | Ref |
| **Sex of child** |  |  |  |  |  |  |  |  |  |
| Male | 210 (49.8) |  | 26 (47.3) | 184 (50.1) | 0.692 |  | 20 (54.1) | 190 (49.4) | 0.585 |
| Female | 212 (50.2) |  | 29 (52.7) | 183 (49.9) | Ref |  | 17 (45.9) | 195 (50.6) | Ref |
| **Maternal knowledge** |  |  |  |  |  |  |  |  |  |
| Inadequate | 124 (29.4) |  | 29 (52.7) | 95 (25.9) | < 0.001^*^ |  | 19 (51.3) | 105 (27.3) | 0.002^*^ |
| Adequate | 298 (70.6) |  | 26 (47.3) | 272 (74.1) | Ref |  | 18 (48.7) | 280 (72.7) | Ref |

| **Characteristics** | **Frequency distribution** |  | **LBW: Low birthweight** | | |  | **SGA: Small for gestational age** | | |
| --- | --- | --- | --- | --- | --- | --- | --- | --- | --- |
|  | **Total (%)**  **n = 422** |  | **LBW (%)**  **n = 55** | **No LBW (%)**  **n = 367** | **p-value ^a^** |  | **SGA (%)**  **n = 37** | **No SGA (%)**  **n = 385** | **p-value ^a^** |
| **Maternal BMI** |  |  |  |  |  |  |  |  |  |
| Underweight | 13 (3.1) |  | 2 (3.7) | 11 (3.0) | 0.798 |  | 1 (2.7) | 12 (3.1) | 0.889 |
| Normal BMI | 279 (66.1) |  | 40 (72.7) | 239 (65.1) | Ref |  | 28 (75.7) | 251 (65.2) | Ref |
| Overweight | 130 (30.8) |  | 13 (23.6) | 117 (31.9) | 0.217 |  | 8 (21.6) | 122 (31.7) | 0.205 |
| **Number of ANC visits** |  |  |  |  |  |  |  |  |  |
| Less than 8 | 304 (72.0) |  | 50 (90.9) | 254 (69.2) | 0.001^*^ |  | 33 (89.2) | 271 (70.4) | 0.015^*^ |
| 8 or more | 118 (28.0) |  | 5 (9.1) | 113 (30.8) | Ref |  | 4 (10.8) | 114 (29.6) | Ref |
| **Number of SP intake** |  |  |  |  |  |  |  |  |  |
| None | 25 (5.9) |  | 2 (3.6) | 23 (6.3) | 0.441 |  | 2 (5.4) | 23 (6.0) | 0.889 |
| 1 – 3 doses | 249 (59.0) |  | 41 (74.6) | 208 (56.7) | Ref |  | 28 (75.7) | 221 (57.4) | Ref |
| > 3 doses | 148 (35.1) |  | 12 (21.8) | 136 (37.0) | 0.027^*^ |  | 7 (18.9) | 141 (36.6) | 0.031^*^ |
| **IFA supplementation** |  |  |  |  |  |  |  |  |  |
| No | 7 (1.7) |  | 0 (0.0) | 7 (1.9) | 0.602 |  | 0 (0.0) | 7 (1.8) | 0.408 |
| Yes | 415 (98.3) |  | 55 (100.0) | 360 (98.1) | Ref |  | 37 (100) | 378 (98.2) | Ref |
| **TD immunization** |  |  |  |  |  |  |  |  |  |
| Not immunized | 31 (7.4) |  | 3 (5.5) | 28 (7.6) | 0.783 |  | 2 (5.4) | 29 (7.5) | 0.636 |
| Immunized | 391 (92.6) |  | 94 (94.5) | 339 (92.4) | Ref |  | 35 (94.6) | 356 (92.5) | Ref |
| **Family planning use** |  |  |  |  |  |  |  |  |  |
| No FP use | 347 (82.2) |  | 47 (85.4) | 300 (81.7) | 0.502 |  | 31 (83.8) | 316 (82.1) | 0.795 |
| FP use | 75 (17.8) |  | 8 (14.6) | 67 (18.3) | Ref |  | 6 (16.2) | 69 (17.9) | Ref |
| **ITNs use** |  |  |  |  |  |  |  |  |  |
| No ITNs use | 152 (36.0) |  | 28 (50.9) | 124 (33.8) | 0.014^*^ |  | 20 (54.1) | 132 (34.3) | 0.017^*^ |
| ITNs use | 270 (64.0) |  | 27 (49.1) | 243 (66.2) | Ref |  | 17 (45.9) | 253 (65.7) | Ref |
| **Malaria infection during pregnancy** |  |  |  |  |  |  |  |  |  |
| Yes | 59 (14.0) |  | 7 (12.7) | 52 (14.2) | 0.774 |  | 5 (13.5) | 54 (14.0) | 0.932 |
| No | 363 (86.0) |  | 48 (87.3) | 315 (85.8) | Ref |  | 32 (86.5) | 331 (86.0) | Ref |
| **Number of pregnancies** |  |  |  |  |  |  |  |  |  |
| 0 – 1 | 111 (26.3) |  | 20 (36.4) | 91 (24.8) | 0.069 |  | 13 (35.1) | 98 (25.5) | 0.201 |
| 2 or more | 311 (73.7) |  | 35 (63.6) | 276 (75.2) | Ref |  | 24 (64.9) | 287 (74.5) | Ref |
| **Number of deliveries** |  |  |  |  |  |  |  |  |  |
| 0 – 1 | 120 (26.3) |  | 21 (38.2) | 99 (27.0) | 0.086 |  | 13 (35.1) | 107 (27.8) | 0.344 |
| 2 or more | 302 (71.6) |  | 34 (61.8) | 268 (73.0) | Ref |  | 24 (64.9) | 278 (72.2) | Ref |
| **Type (place) of delivery** |  |  |  |  |  |  |  |  |  |
| Skilled (facility) | 392 (92.9) |  | 50 (90.9) | 342 (93.2) | 0.540 |  | 33 (89.2) | 359 (93.2) | 0.359 |
| Unskilled (at home) | 30 (7.1) |  | 5 (9.1) | 25 (6.8) | Ref |  | 4 (10.8) | 26 (6.8) | Ref |
| **Mode of delivery** |  |  |  |  |  |  |  |  |  |
| Vaginal | 379 (89.8) |  | 48 (87.3) | 331 (90.2) | 0.505 |  | 31 (83.8) | 348 (90.4) | 0.205 |
| Caesarian section | 43 (10.2) |  | 7 (12.7) | 36 (9.8) | Ref |  | 6 (16.2) | 37 (9.6) | Ref |

| **Hb levels across trimesters** | **Frequency distribution** |  | **LBW: Low birthweight** | | |  | **SGA: Small for gestational age** | | |
| --- | --- | --- | --- | --- | --- | --- | --- | --- | --- |
|  | **Total (%)** |  | **LBW** | **No LBW** | **p-value ^a^** |  | **SGA** | **No SGA** | **p-value ^a^** |
| **First-trimester Hb levels** |  |  |  |  |  |  |  |  |  |
| Anaemia | 268 (63.5) |  | 38 (69.1) | 230 (62.7) | 0.356 |  | 25 (67.6) | 243 (63.1) | 0.591 |
| Normal | 129 (30.6) |  | 10 (18.2) | 119 (32.4) | Ref |  | 6 (16.2) | 123 (32.0) | Ref |
| Polycythaemia | 25 (5.9) |  | 7 (12.7) | 18 (4.9) | 0.022^*^ |  | 6 (16.2) | 19 (4.9) | 0.005^*^ |
| **Second-trimester Hb levels** |  |  |  |  |  |  |  |  |  |
| Anaemia | 301 (71.3) |  | 45 (81.8) | 256 (69.7) | 0.065 |  | 28 (75.7) | 273 (70.9) | 0.540 |
| Normal | 106 (25.1) |  | 2 (3.6) | 104 (28.3) | Ref |  | 1 (2.7) | 105 (27.3) | Ref |
| Polycythaemia | 15 (3.6) |  | 8 (14.6) | 7 (1.9) | < 0.001^*^ |  | 8 (21.6) | 7 (1.8) | < 0.001^*^ |
| **Third-trimester Hb levels** |  |  |  |  |  |  |  |  |  |
| Anaemia | 191 (45.3) |  | 51 (92.7) | 140 (38.1) | < 0.001^*^ |  | 33 (89.2) | 158 (41.0) | < 0.001^*^ |
| Normal | 224 (53.0) |  | 1 (1.8) | 223 (60.8) | Ref |  | 1 (2.7) | 223 (57.9) | Ref |
| Polycythaemia | 7 (1.7) |  | 3 (5.5) | 4 (1.1) | 0.050 |  | 3 (8.1) | 4 (1.0) | 0.001^*^ |
